# Polygenic regulation of PTSD severity and outcomes among World Trade Center responders

**DOI:** 10.1101/2020.12.06.20244772

**Authors:** Laura M. Huckins, Jessica S. Johnson, Leo Cancelmo, Olivia Diab, Jamie Schaffer, Leah Cahn, Cindy Aaronson, Sarah R Horn, Clyde Schechter, Shelby Marchese, Linda M Bierer, Iouri Makotkine, Frank Desarnaud, Janine D Flory, Michael Crane, Jacqueline M. Moline, Iris G. Udasin, Denise J. Harrison, Panos Roussos, Dennis S. Charney, Guia Guffanti, Karestan C Koenen, Rachel Yehuda, Steven M. Southwick, Robert H. Pietrzak, Adriana Feder

## Abstract

Post-traumatic stress disorder (PTSD) is a debilitating psychiatric condition triggered by exposure to trauma. The study of PTSD is complicated by highly heterogeneous presentations and experiences of trauma between individuals. Capitalizing on the existence of the World Trade Center General Responder Cohort (WTC-GRC) of rescue, recovery and clean-up workers who responded during and in the aftermath of the World Trade Center (WTC) 9/11/2001 attacks, we studied genetic correlates of PTSD in a sample of 371 WTC responders, selected from the WTC-GRC utilizing stratified random sampling. This deeply phenotyped sample of WTC responders – ranging from no/low PTSD symptom levels to severe PTSD– provide a unique opportunity to study genetic risk factors for PTSD severity and chronicity following a single, shared, well-documented trauma, also incorporating measures of childhood and other lifetime traumas.

We examined associations of polygenic risk scores (PRS) –derived from a range of genome-wide association studies (GWAS) of behavioral traits, psychiatric disorders, and brain volumetric phenotypes– with PTSD severity and chronicity among these 371 individuals. Our results demonstrate significant genetic regulation of lifetime PTSD severity, assessed with the lifetime version Clinician-Administered PTSD Scale (CAPS), and chronicity, assessed with the past-month CAPS. PRS derived from GWAS of attention deficit-hyperactivity disorder (ADHD), autism spectrum disorder (ASD), and brain imaging phenotypes (amygdala and putamen volumes) were associated with several PTSD symptom dimensions. Interestingly, we found greater genetic contribution to PTSD among cases compared to our full sample. In addition, we tested for associations between exposures to traumatic stressors, including WTC-related exposures, childhood trauma, and other lifetime traumatic life events in our full sample. Together, polygenic risk and exposures to traumatic stress explained ~45% of variance in lifetime CAPS (R^2^=0.454), and ~48% of variance in past-month CAPS (R^2^=0.480) in the full sample.

These participants represent a highly vulnerable population, with exposures to severe trauma during 9/11 and the following days and months. These novel identified associations between PTSD and PRS of behavioral traits and brain volume phenotypes, as well as replicated associations with PRS of other psychiatric disorders, may contribute to understanding the biological factors associated with risk for and chronicity of PTSD. In particular, the identification of neuroimaging phenotypes indicates that coupling of neuroimaging with genetic risk score calculations may predict PTSD outcomes.

## Introduction

Understanding the biological factors contributing to vulnerability to PTSD will be key in developing effective treatments for the disorder, and in identifying individuals who may be at high risk of development and persistence of PTSD. In line with this goal, twin studies have established substantial genetic aetiology of PTSD, with heritability estimates around 30%^1–5^, although these vary substantially according to sex and trauma type^1,3,5–7^. Significant progress in elucidating the genetic aetiology of PTSD has been made through the advent of large-scale genome-wide association studies (GWAS). Collaborative efforts from the PTSD working group of the Psychiatric Genomics Consortium (PGC) established single nucleotide polymorphism (SNP)-heritability at 5-20%, again with heritability estimates varying by sex^2^. Progress in identifying significant GWAS loci and other biomarkers is likely hampered by heterogeneity within our studies. Deeper insights into PTSD biology may be gained by leveraging small but deeply phenotyped cohorts. In particular, identifying single, quantifiable traumatic events is rare.

The worker response to the World Trade Center (WTC) disaster was unprecedented in scope, involving tens of thousands of traditional and non-traditional responders in rescue, recovery and clean-up efforts. PTSD arising after equally unprecedented traumatic exposures remains highly prevalent (10-22%) and persistent in this population in the second decade following 9/11^8–10^. Here, we characterized a unique sample of 371 WTC responders of diverse backgrounds and training, all exposed to the aftermath of the 9/11 terrorist attacks, recruited on average 13.5 years after 9/11/2001 and ranging in PTSD symptom severity from no/low symptoms to severe PTSD. The present study aimed to fill a major gap in our understanding of the development and chronicity of this disabling condition by conducting genome-wide genetic and gene expression profiling^11^ in a representative sample of WTC responders, characterized in depth with in-person psychiatric, medical, and psychosocial assessments. The existence of this cohort represents a unique opportunity to study genetic risk factors for PTSD following a shared and well-documented trauma in a sample of individuals who nonetheless vary greatly in severity of exposures to WTC-related trauma and other lifetime traumas.

Although few genome-wide associations have been identified for PTSD to date, genetic aetiology and polygenicity of the disorder is well established; that is, small, additive effects across the genome combine to increase risk for PTSD development after trauma exposure. Here, we tested whether a range of polygenic risk scores (PRS) derived from GWAS of psychiatric disorders and behavioral traits, and from available GWAS of magnetic resonance imaging (MRI) phenotypes relevant to PTSD, predict PTSD diagnosis, severity and chronicity within our sample. First, we tested whether polygenic risk predicts WTC-related PTSD severity, using the total score on the Clinician Administered PTSD Scale (CAPS), lifetime version, as a proxy for highest severity of WTC-related PTSD symptoms since 9/11/2001, among the full sample, and among cases only. Next, we tested whether the total past-month CAPS score is also genetically regulated among our cohort. Given the substantial time elapsed (on average 13.5 years) since the WTC-related trauma exposures, this score served as a measure of chronicity of WTC-related PTSD symptoms. The depth of phenotyping and characterization of this cohort also enabled us to test the contribution of lifetime trauma exposures and other stressful life events to WTC-related PTSD severity and chronicity (including exposures during childhood, 9/11 and its aftermath, subsequent stressful life events, and other lifetime traumas), both individually and in concert. Finally, we tested whether polygenic risk and potentially traumatic exposures interact to increase PTSD risk and severity. Examining this interplay between genetic and exposure variables, and understanding their individual and synergistic contributions to PTSD development and chronicity will be vital to elucidating biological mechanisms underlying the disorder.

## Methods

### Participants

The WTC Health Program (WTC-HP) is a regional consortium of five clinical centers established in the greater New York City area by the Centers of Disease Control and Prevention in 2002, with the goal of providing health monitoring and treatment to WTC responders, comprising the WTC-HP General Responder Cohort^12^. In earlier studies in this cohort, we characterized the severity, chronicity, and psychosocial predictors and correlates of PTSD symptoms using data from self-report questionnaires^9,13,14^. For the present study, we recruited participants from the WTC-HP Responder Cohort who had completed at least three periodic health monitoring visits at one of the four WTC-HP clinical centers participating in this study – Mount Sinai Medical Center, New York University, Northwell Health, and Rutgers/The State University of New Jersey – and who had provided signed consent to be contacted for research studies. Stratified random sampling was employed to ensure selection of WTC responders spanning the full range of WTC-related PTSD symptom severity, from no/minimal symptoms to severe/chronic PTSD symptom levels on the PTSD Checklist – Specific Version (PCL-S)^15^ completed during periodic health monitoring visits to the WTC-HP. We additionally enriched our sample with WTC responders from the two “extreme groups”, by oversampling individuals with persistently high PTSD symptom levels and those with no/minimal PTSD symptoms, based on PCL-S scores from their first three WTC-HP periodic health monitoring visits. Individuals with a lifetime history of chronic psychotic disorder or bipolar disorder type I, substance abuse/dependence or alcohol dependence over the prior three months, current pregnancy, acute medical illness or exacerbation of chronic medical illness, history of significant head injury or cerebrovascular accident, changes in medications or medication dosages over the prior month, or who were taking oral or regularly injected steroid medications were excluded from the study.

The study, conducted between April 2013 and September 2017, was approved by the Institutional Review Board of the Icahn School of Medicine at Mount Sinai, and all participants provided written informed consent. A total of 471 WTC responders completed in-person clinical assessments, yielding a final sample of 371 participants who met study eligibility criteria and completed study procedures. Mean age (SD) was 54.2 (8.4) years, and 82.5% were male. The sample was composed of 41.2% police responders and 58.8% non-traditional responders (e.g., construction workers).

### Assessments

Data on 10 WTC-related exposures (e.g., exposed to human remains, received treatment for an illness/injury during WTC recovery work) was obtained from interviews and self-report questionnaires completed by participants during their first health-monitoring visit to the WTC-HP, an average of 4.3 (SD=2.7) years following 9/11/2001, yielding the WTC Exposure Index (total count of exposures, range 0-10). In-person clinical assessments were conducted an average of 13.5 (SD=1.3) years following 9/11/2011. Trained Masters- or PhD-level clinical interviewers administered the Structured Clinical Interview for DSM-IV^16^ (SCID) to assess current and lifetime Axis-I psychiatric diagnoses, and the Clinician-Administered PTSD Scale^17^ (CAPS), lifetime and past-month versions, to assess lifetime and past-month WTC-related PTSD symptom severity and WTC-related PTSD diagnostic status. Lifetime and past-month PTSD diagnosis was defined as meeting DSM-IV criteria for WTC-related PTSD and a total score ≥ 40 on the lifetime and past-month CAPS, respectively.

On the same day as the clinical assessment, participants also completed the Childhood Trauma Questionnaire^18^ (CTQ), assessing physical, sexual, and emotional abuse, and physical and emotional neglect experienced in childhood; the Traumatic Life Events Questionnaire^19^ (TLEQ), assessing lifetime exposure to a range of traumatic events (e.g, crime, natural disaster, assault); a checklist asking which of 15 stressful life events they had experienced since 9/11/2011 (e.g., “lost a job/laid off/lost income”, “divorced from spouse”, “had debt trouble”), modified from the Diagnostic Interview Schedule (DIS) Disaster Supplement^20^; and a health questionnaire asking which medical conditions they had ever been diagnosed with^21^, modified to add common WTC-related conditions (asthma or chronic respiratory condition, chronic rhinitis or sinusitis, sleep apnea, or acid reflux). Participants additionally completed a history and physical examination conducted by a licensed nurse practitioner, as well as routine laboratory testing, to rule out acute medical illness or exacerbation of chronic medical illness.

### Blood Sample Collection and Genotyping

Participants underwent collection of fasting blood samples approximately between 8:00 and 9:00 am. Genomic DNA was purified from an aliquot of 200 microliters of whole blood collected in PAXgene blood DNA tubes (Qiagen, Germantown, MD, USA) using a QuickGene DNA whole blood Kit S (Kurabo, Osaka, Japan) followed by an automated DNA extraction using a QuickGene-810 instrument according to the manufacturer instructions (Kurabo, Osaka, Japan).

Genomic DNA quality and concentration were estimated using a NanoDrop 200c spectrophotometer according to the manufacturer instructions (Thermo Fisher Scientific, Waltham, MA, USA). Samples with an optical density ratio 260/280 superior or equal to 1.8 passed the quality control.

### Genotyping Quality Control

Samples were genotyped in two batches (195 in the first, and 186 in the second). The following QC steps were applied separately to each batch. Standard genotyping quality control metrics were applied using plink^22,23^ to remove: multi-allelic variants, indels, ambiguous SNPs (c/g or a/t); variants failing Hardy-Weinberg Equilibrium expectations (HWE<1e-06); variants with MAF<0.1%; individuals with high missingness (>0.05) or genotypes with low call-rate (>0.05)

Following both individual- and variant-level QC, genotypes were imputed using the Michigan Imputation server^24^. Genotypes were imputed using the 1000 Genomes Phase 3 (Version 5) reference panel^25,26^, and standard imputation QC performed according to Michigan Imputation server guidelines^24^. Following imputation, a final variant-level QC was performed to remove variants with low imputation quality scores (R<0.2), and to repeat previous HWE, MAF, call-rate and missingness filters.

Following genotype-level QC, we merged the two batches, retained overlapping variants only, and repeated our MAF, HWE, call-rate and genotyping filters described previously on the full, merged cohort. In total, we retained 381 individuals and 10,858,359 variants. We computed relatedness and genotype-derived ancestry using plink^22,23^. We identified one pair of samples with high relatedness (pihat=0.55), and removed one of the pair at random. In total, 371 individuals remained following our genotype and sample level QC steps.

### Genetic Regulation of PTSD

Polygenic risk scores (PRS) for psychiatric diagnoses, personality traits, and MRI brain volume-related phenotypes were calculated using 28 publicly available GWAS summary statistics (**Table 1**). Briefly, PRS are calculated for each individual within a cohort as a weighted sum of risk alleles for a given trait or disorder. For example, a schizophrenia-PRS is calculated as a sum of schizophrenia risk alleles in each individual’s genome, weighted by the effect size of each variant on schizophrenia risk. For each trait, we tested for association with (1) total lifetime (highest) CAPS score and (2) total past-month CAPS score using PRSice^27,28^, including all significant genotype-derived principal components to control for confounding due to ancestry. The amount of phenotypic variance explained by the GWAS-derived PRS alone was estimated using a nested approach (equation 1).

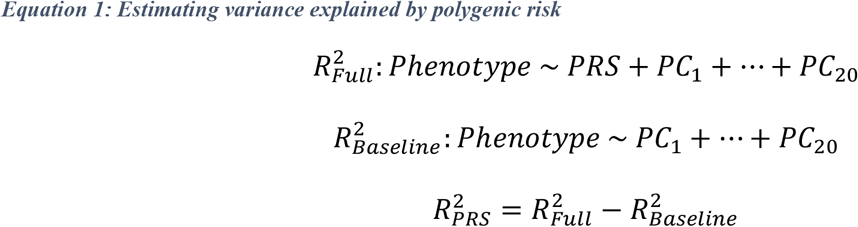

Given that a large number of individuals within our cohort have low CAPS scores, our associations may be driven by this ‘control’ part of our distribution. Therefore, we repeated our analysis, restricting only to ‘cases’: i.e., individuals meeting the CAPS and PTSD diagnosis requirements described in our methods section.

For all significant PRS, we tested for interaction effects between polygenic risk and sex in our full cohort.Given the high degree of shared genetic heritability between psychiatric diagnoses, traits, and brain-volume related phenotypes, modelling these PRS separately likely introduces redundancy, and may over-estimate the amount of phenotypic variance explained by PRS Therefore, we next constructed a joint model, including all PRS reaching p<0.1 in our individual models. We calculated PRS for each trait at ten standard p-value thresholds (0.0001, 0.001, 0.01, 0.05, 0.1, 0.2, 0.3, 0.4, 0.5, 1), and fit a joint model at each threshold. We adjusted for genotype-derived principal components using a nested approach as previously.

### Accounting for Trauma Exposure

The samples included in this study have deep phenotyping information, including details of experiences of exposures during 9/11 and its aftermath, as well as stressful life events since 9/11 and other lifetime traumas. These exposure variables may provide vital insights into modifying effects of environment on genetic predisposition.

First, we tested whether WTC-related and other lifetime trauma exposures, as well as stressful life events since 9/11/2001, significantly predicted WTC-related PTSD outcome. Specifically, we tested for association of WTC Exposure Index, total CTQ and TLEQ scores, and total count of stressful life events since 9/11/2001 with (i) total lifetime CAPS and (ii) total past-month CAPS scores. We tested for each WTC Exposure Index item individually, as well as for total WTC Exposure Index, total CTQ, and total TLEQ score, including sex, age, and the first 20 genotype-derived principal components as covariates. In addition, we tested for significant interaction effects between responder type (police vs. non-traditional responder) and with each of the ten WTC Exposure Index items individually. We estimated total variance in CAPS scores explained using a nested model (equation 2). Baseline factors (sex, age, principal components) predicted ~6% of variance in CAPS scores among the full cohort (R^2^= 0.06277, 0.05215 in lifetime and past-month CAPS scores, respectively).

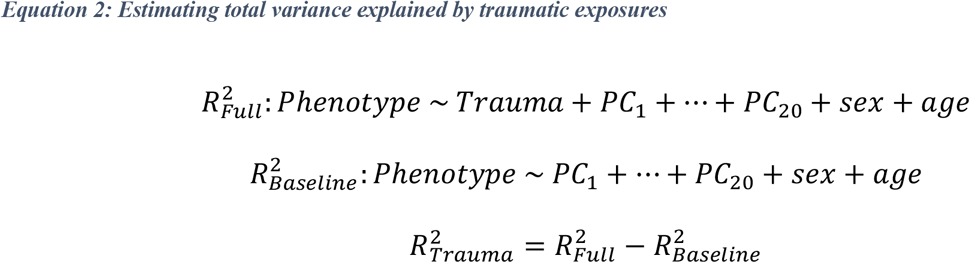

Similar to our PRS model, we expect some overlap between these different exposures and factors; therefore, we fit a joint model to identify factors most significantly predicting CAPS scores. Finally, we fit a joint model including all individual exposures and PRS in order to predict (i) lifetime (highest) and (ii) past-month CAPS.

## Results

### Characterization of PTSD among 371 WTC responders

All responders in the final study sample of 371 completed the CAPS lifetime and past-month versions, CTQ, TLEQ, and the checklist about stressful life events experienced since 9/11/2011, modified from the DIS Disaster Supplement. The WTC Exposure Index was computed from participants’ responses obtained at their first WTC-HP health monitoring visit. A total of 112 (29.9%) responders met diagnostic criteria for lifetime WTC-related PTSD (**Figure 2A**; 71 non-traditional and 41 police responders). Forty-three (11.5%) met diagnostic criteria for past-month PTSD (**Figure 2B**; 33 non-traditional and 10 police responders).

**Figure 1:**
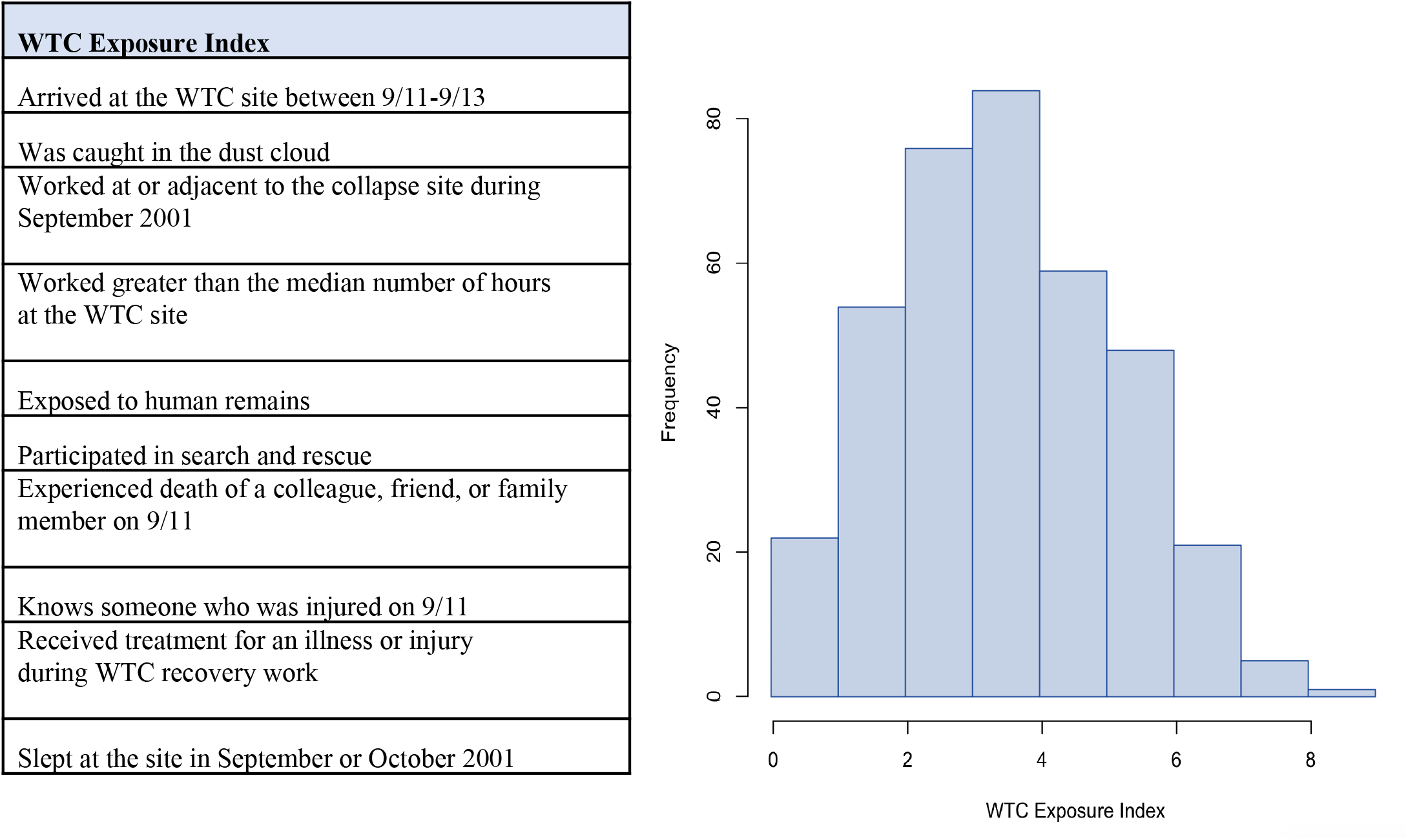
World Trade Centre Exposures were quantified using the World Trade Center Exposure Index. (A) WTC Exposure Index Factors. (B) Distribution of total WTC Exposure Scores.

**Figure 2:**
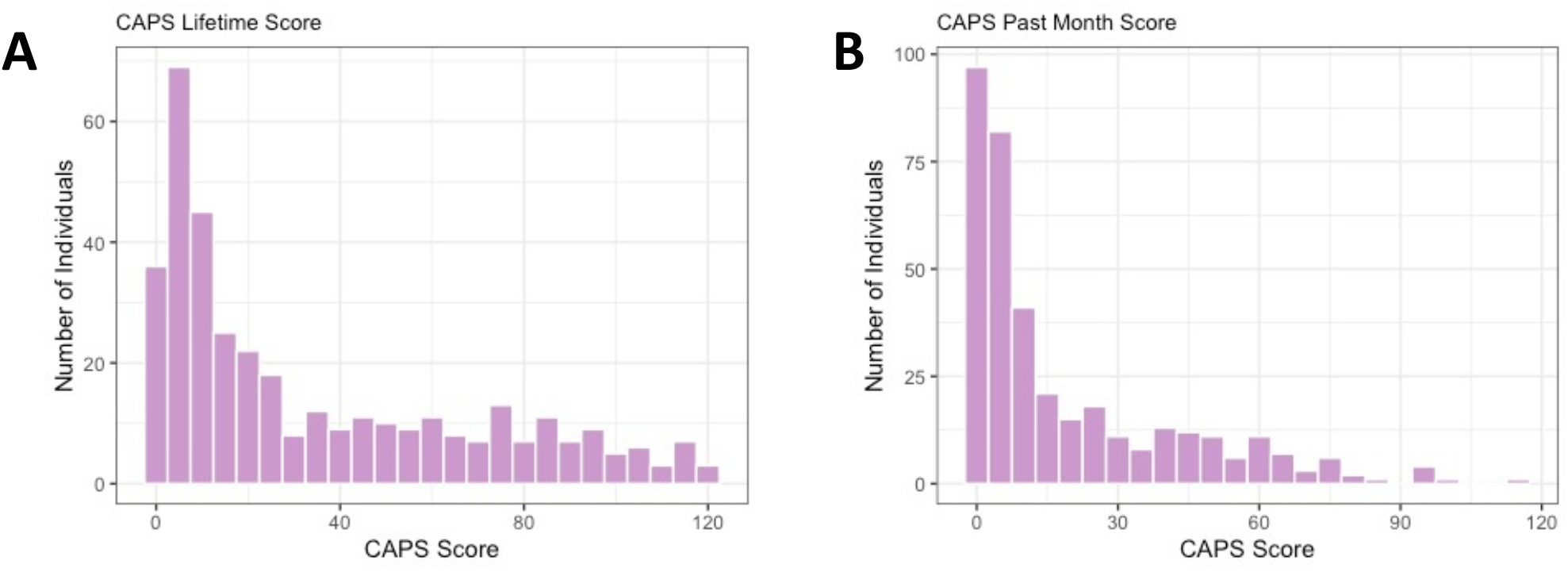
Characterization of PTSD among 371 WTC responders. A) Highest recorded lifetime CAPS score; B) Past-month CAPS score

### Polygenic Risk Scores predict PTSD outcomes and severity among WTC responders

We performed PRS analyses using 28 GWAS for psychiatric disorders, traits, and MRI-derived brain measures, and tested ability to predict lifetime and past-month CAPS scores. Risk scores derived from GWAS of putamen and amygdala volumes, attention deficit-hyperactivity disorder (ADHD), anxiety, autism spectrum disorder (ASD), smoking behavior (“ever smoker”), self-defined wellbeing, and bipolar disorder all significantly predicted lifetime CAPS scores (R^2^>0.01; p<0.047; **Table 1**); the same traits (except self-defined wellbeing and smoking) also predicted past-month CAPS score (R^2^>0.012; p<0.030; **Table 1**). In addition, PTSD-PRS derived from the latest PGC-PTSD GWAS significantly predicted past-month CAPS score (R^2^=0.017, p=0.012), although PTSD-PRS did not significantly predict lifetime CAPS (R^2^ =0.0094, p=0.06).

Next, we repeated our analysis including only PTSD ‘cases’, i.e., individuals meeting DSM-IV criteria for WTC-related PTSD and with a total CAPS score ≥ 40. We repeated this analysis for lifetime cases, and past-month cases, following these definitions. We identified significant associations between lifetime (highest) CAPS score and PRS derived from GWAS of anxiety, mean putamen and amygdala volume, smoking behavior, and schizophrenia (R^2^>0.042; p<0.045), implying that these PRS predict PTSD severity (**Table 2**). In addition, PRS from GWAS of schizophrenia, loneliness, mean putamen and amygdala volumes, smoking behaviour, and ASD all predicted past-month CAPS score among cases (R^2^>0.044; p<0.033).

Finally, we compared the proportion of variance explained by PRS in the full cohort vs. cases only, and found that substantially more variance was explained among cases (**Figure 3**), implying that PTSD severity is more significantly genetically regulated than PTSD case/control status.

**Figure 3:**
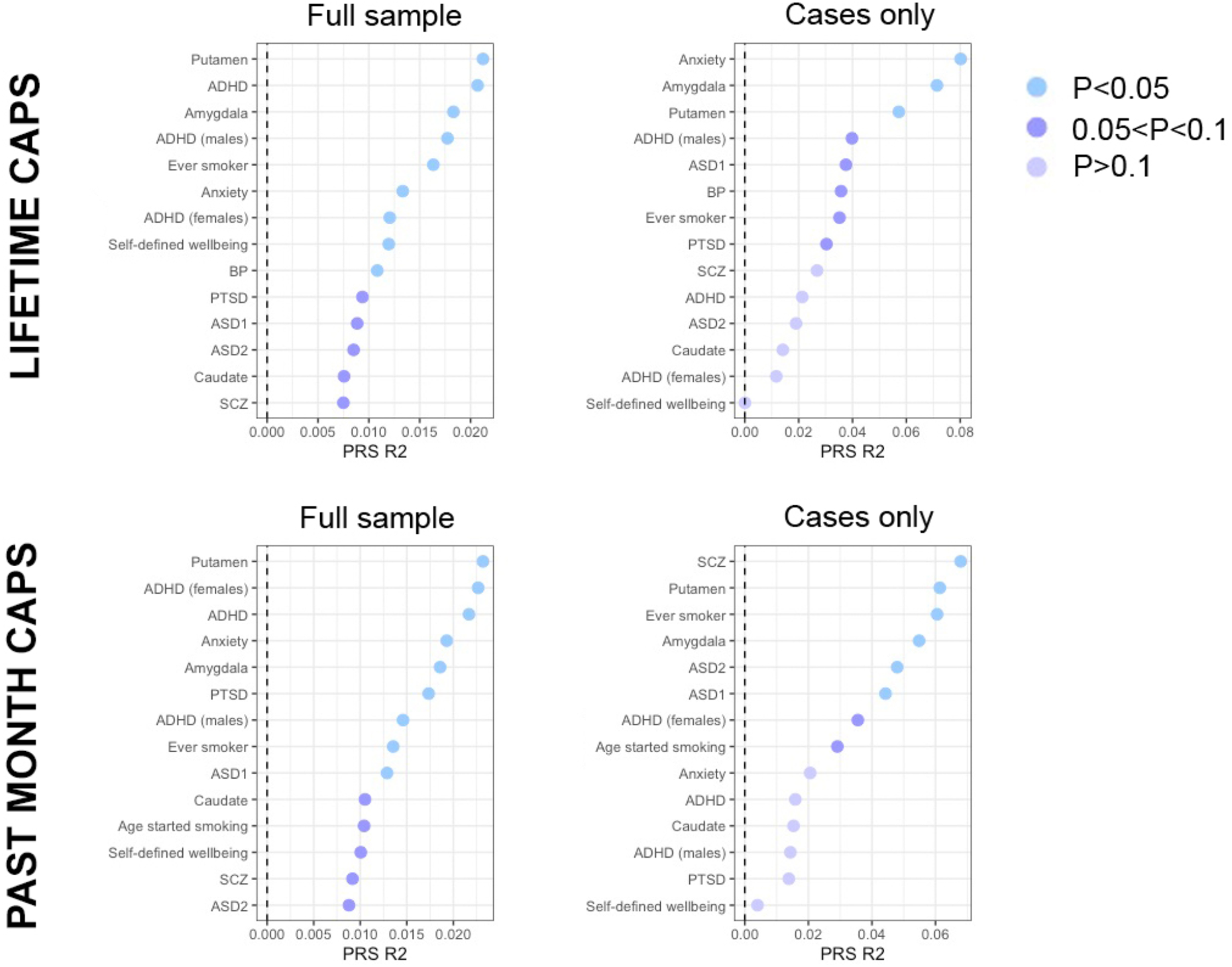
Phenotypic variance (R^2^) in (A) Highest Lifetime and (B) Past Month CAPS explained by PRS. All traits with p<0.1 are shown here for (Left) Full sample and (Right) among cases only.

### Polygenic Risk Scores jointly explain significant variance in total CAPS score

We fit a joint PRS model, including all GWAS-derived PRS reaching p<0.1 in in the individual analyses simultaneously. In the full cohort, the final model explained 6.7% of variance in total lifetime (highest) CAPS score (R^2^=0.0670; most significant PRS: mean amygdala volume, ADHD, self-defined wellbeing; **Table 3**), and 6.5% of variance in total past-month CAPS score (R^2^=0.0654; most significant PRS: mean putamen volume, anxiety). Repeating this analysis among cases only revealed a significant increase in proportion of variance explained (lifetime CAPS score R^2^=0.21, most significant PRS: anxiety, alcohol dependence; past-month CAPS R^2^=0.157, most significant PRS: mean putamen volume, ever smoker), implying that severity of PTSD may be more significantly genetically regulated than PTSD case/control status.

### Characterization of PTSD and potentially traumatic exposures

WTC-related exposure severity during 9/11 and its aftermath, operationalized with the WTC Exposure Index, significantly predicted both total lifetime (highest) and past-month CAPS scores (p=9.82×10^−5^, 1.25×10^−4^ respectively; **Table 4A**). In addition, three specific exposures within the WTC Exposure Index significantly correlated with current PTSD symptom severity; “Knowing someone who was injured on 9/11” (p=0.0011, p=8.55×10^−4^ for lifetime and past-month CAPS scores, respectively), “Received treatment for an injury or illness during WTC recovery work” (p=8.81×10^−6^, p=6.74×10^−6^), and “Slept on-site during September or October2001” (p=0.0037, p=0.011). In addition, knowing someone injured on 9/11 significantly predicted highest lifetime CAPS score among cases alone (p=0.033). No specific WTC-related exposures predicted past-month CAPS score among cases.

Next, we tested for interaction effects between WTC exposures and responder type (police vs. non-traditional responder). We found a significant positive interaction between responder type and “Knowing someone who was injured on 9/11” (β=17.2, p=0.024) on lifetime (highest) CAPS score, indicating that this effect is stronger among non-traditional responders; i.e., knowing someone injured on 9/11 had a greater effect on CAPS among non-traditional responders. Conversely, we found a significant negative correlation between responder type and “Received treatment for an injury or illness during WTC recovery work” on past-month CAPS score (β=-13.0, p=0.0044), indicating that this effect is stronger among police responders; i.e., receiving treatment for an injury or illness had a greater effect on CAPS among police responders than among non-traditional responders.

Modelling all WTC exposures jointly explained ~25% of variance in total lifetime and past-month CAPS scores (R^2^=0.25, 0.24; p=6.27×10^−13^, 2.585×10^−11^, respectively: **S. Table 1**) among the full sample. However, modelling all WTC exposures did not significantly predict total lifetime (highest) or past-month CAPS scores among cases.

Although the CAPS was administered specifically with regards to WTC exposures, we hypothesize that prior and subsequent traumas and stressful life events may also affect WTC-related PTSD outcomes; therefore, we tested for association with stressful life events since 9/11/2001, as well as childhood trauma severity and lifetime trauma exposures assessed with the CTQ and TLEQ, respectively (**Table 4**). We expect that these various measures of trauma and stressors will include some degree of redundancy, particularly among some of our most significant variables (for example, among CTQ subscale scores). Therefore, we constructed joint models to estimate the proportion of phenotypic variance in (i) lifetime and (ii) past-month CAPS scores explained by all exposures jointly. We were able to significantly predict both lifetime (R^2^=0.44) and past-month (R^2^=0.44) CAPS scores. Although the CAPS was employed to assess WTC-related PTSD symptoms, the most significant exposures associated with both lifetime and past-month CAPS scores were number of stressful life events since 9/11/2001, and childhood physical and emotional neglect. Additional significant predictors of lifetime (highest) CAPS score included responder type, alone and in interaction with WTC-related exposure “Knowing someone who was injured on 9/11”, and total TLEQ score. Interestingly, significant predictors of past-month CAPS score also included a number of specific WTC Exposures, as well overall WTC exposure severity (WTC Exposure Index), and the interaction between responder type and WTC exposure “Received treatment for an injury or illness during WTC recovery work” (**Table 4A**).

### Interactions between Exposures and Polygenic Risk

Finally, we tested for interactions between all significant PRS and exposures. We identified four significant interactions, all predicting past-month CAPS score, between childhood physical neglect and anxiety-PRS (p=0.048), childhood physical neglect and ADHD-PRS (p=0.0085), stressful life events since 9/11/2001 and ever smoker-PRS (p=0.036), and childhood emotional neglect and ever smoker-PRS (p=0.011).

Next, we constructed joint models for lifetime and past-month CAPS, both in our full cohort and among cases only, including in each score all previously significant PRS, exposures and interaction terms between responder type and WTC exposures, and between PRS and exposures (for past-month CAPS only). Our models explained ~45% of variance in lifetime CAPS score among the full cohort (R^2^=0.454), and ~48% of variance in past-month CAPS score (R^2^=0.480) (**S. Table 2**).

## Discussion

Our analysis examined genetic and environmental factors associated with WTC-related PTSD severity and chronicity, assessed with the CAPS during in-person clinician-administered interviews: first, with highest lifetime CAPS score, a measure of highest PTSD severity since 9/11; second, with past-month CAPS, a measure of PTSD chronicity. PRS derived from GWAS of ADHD, ASD, anxiety and brain imaging phenotypes (amygdala and putamen volume), among others, jointly explained ~7% of variance in CAPS scores within our cohort, in line with the degree of phenotypic variance in psychiatric diagnoses observed in previous studies^29^.

Our study identifies a number of novel insights into the genetic architecture of PTSD. First, to our knowledge, this is the first evidence for shared genetic aetiology between PTSD and MRI-derived amygdala and putamen volumes. Importantly, imaging studies have previously identified consistent associations between smaller amygdala volumes and PTSD diagnosis^30,31^, in line with the direction of effect in our study; however, such studies necessarily occur after PTSD onset, making causality and direction of effect difficult to infer. Demonstrating shared genetic aetiology between these traits therefore provides novel and vital context to these existing analyses. Our study is also the first evidence of shared genetic risk factors between PTSD, ADHD, ASD, and self-defined wellbeing. Furthermore, despite substantial evidence for shared genetic aetiology between PTSD and smoking established in twin studies, we believe this is the first study to demonstrate significant shared polygenic risk between the two traits.

In addition to these polygenic associations, our study also provides novel insights into the genetic architecture of PTSD through three key strengths of our study design. First, our analysis focuses on a quantitative trait (CAPS), conferring substantially higher power than traditional case/control designs^32^. Second, this design allows us to probe the role of polygenic risk in PTSD severity, rather than only in predicting PTSD diagnosis. Third, we probe specifically the role of polygenic risk and exposure to potentially traumatic events among cases only. In particular, comparing both individual and joint-PRS approaches revealed that PRS explained significantly more variance in CAPS among cases compared to the full cohort, implying that PTSD severity may be more strongly genetically regulated than PTSD case-control status. In parallel, we observed less significant relationships between traumatic exposures and PTSD severity among cases than in the full population. Together, these findings point towards greater genetic regulation of PTSD severity among cases than among the full population. This finding in particular has significant implications for larger genetic analyses of PTSD, which traditionally focus on case-control status, or on PTSD among the full population, rather than studying PTSD severity among cases specifically.

We additionally found significant associations between a several WTC-related exposures and both highest lifetime and past-month CAPS; these associations differed significantly according to responder type. In particular, we found a more significant effect of “Knowing someone who was injured on 9/11” among non-traditional responders, and a of “Treated for injury or illness during the WTC recovery effort” among police responders. These findings might in part reflect higher severity and persistent personal impact of WTC-related exposures involving injury or illness^33,34^, which might differ by responder type. Of note, prior research in the larger WTC responder cohort found an association between WTC-related exposure severity and WTC-related PTSD symptom severity specifically in police responders^35^.

Importantly, despite the focus of our study specifically on WTC-related PTSD, as ascertained in clinician interviews, the most significant factors predicting both highest lifetime and past-month PTSD were “experiencing new life stressors since 9/11”, and childhood physical and emotional neglect scores, rather than WTC-related exposures, the latter explaining substantially less phenotypic variance in CAPS scores. These findings are in line with those of prior research on the impact of both childhood neglect^36^ and secondary stressors^34^ on risk for PTSD, and have significant implications for future studies of PTSD. We, along with others, have shown that the genetic aetiology of PTSD differs significantly according to trauma type^5^ and gender^37,38^, implying that stratifying by trauma type may improve study power and increase biological insights available from existing studies. We further suggest that studies focusing on the role of traumas in PTSD, or seeking to maximize power of genome-wide association studies, should seek to characterize a range of exposures to traumas and stressful life events, rather than assuming a single causal or index trauma.

Within our full cohort, significantly more variance in highest lifetime and past-month CAPS was explained by reported traumas and stressors than by genetic factors. To some extent, this likely reflects the relatively low power of many of the GWAS from which we derive our PRS, and limitations in applying PRS to individuals not of solely European ancestry. Moreover, we note that, to date, no large-scale GWAS of CAPS are available for inclusion in our PRS analysis, and we therefore derived our PTSD-PRS from a large PTSD case/control GWAS^38^. Although ostensibly studying the same underlying phenotype (PTSD), this difference in definitions may lead to reduced predictive power in our analysis. In line with this, we find that the PTSD-PRS significantly predicts CAPS in our full sample, but not among cases only, perhaps suggesting that the PTSD-PRS captures genetic predisposition to PTSD rather than PTSD severity. Consequently, we may be significantly *under-estimating* the amount of variance explained by genetic risk factors. Simultaneously, we may significantly *over-estimate* the amount of variance explained by these traumas and exposures to traumatic stress. Most importantly, participant recall of traumas and stressors is biased by current mood and PTSD symptomatology^39,40^, implying that individuals within our study with current PTSD may rank their stressors higher than those without, confounding our analysis. The effect may be especially pronounced among scales and scoring systems that require rankings or subjective recall many years later (for example, the CTQ or ‘stressful life events since 9/11’) than the data on WTC-related exposures collected within a few years post-9/11/2001, perhaps also accounting for the difference in variance explained by these factors^41^.

Finally, although our joint PRS-trauma exposure models explained roughly similar amounts of variance in lifetime and past-month CAPS scores, we note strikingly different contributions in each model. In particular, we note that more variance in lifetime highest CAPS score than in past-month CAPS score is explained by genetic factors, which may reflect differential genetic regulation of the two phenotypes (i.e., that PTSD severity is more strongly genetically regulated than PTSD chronicity). In addition, we note substantial differences in the stressors and traumas associated with each phenotype, with many WTC-related exposures significantly associated with past-month CAPS only. Together, these findings imply different genetic architectures and environmental stressors contributing to these two phenotypes.

In contrast to previous studies, which identified sex-specific genetic aetiology in PTSD, we did not observe any significant interactions between sex and polygenic risk, although this is likely attributable to the small number of women in our sample. To some extent, the proportion of variance explained, and the lead GWAS-PRS associations identified in our analysis may be due to the power of each individual GWAS, rather than due to true, shared biology. For many of the GWAS from which we derive our polygenic-risk scores, we likely capture only a fraction of the true SNP-based heritability in psychiatric disease in current studies^37^, and we expect an effect which may be more severe among studies of those phenotypes which are of particular interest to us: for example, PTSD and MDD. Similarly, the significant associations that we observe in our single-PRS analysis for schizophrenia and bipolar disorder may reflect the substantial power of those GWAS studies, rather than necessarily indicating that these disorders have more shared genetic aetiology with PTSD than, for example, MDD.

These participants represent a highly vulnerable population, with exposures to severe trauma during 9/11 and its aftermath. Identifying individuals at heightened risk for PTSD, or increased risk for severe or chronic PTSD, is of special importance and relevance in this group. Our study leverages a powerful quantitative trait definition (CAPS), together with deep phenotyping to yield novel insights into the genetic architecture of PTSD and PTSD severity. In particular, our analysis identifies novel associations between PTSD and ADHD, ASD, self-defined wellbeing, and smoking behaviours. Furthermore, we identify shared polygenic risk between PTSD and MRI imaging phenotypes, indicating that coupling of neuroimaging with genetic risk score calculations may predict PTSD outcomes and resilience.

## Supporting information

Main Tables

## Data Availability

Summary data will be made available upon publication.

## Acknowledgements

We are grateful for the advice and guidance of Pamela Sklar, who contributed centrally to the design of this study.

## Notes

### Competing Interest Statement

The authors have declared no competing interest.

### Funding Statement

This work was supported by NIOSH: Biomarkers of Psychological Risk and Resilience in WTC Responders
U01 OH010407
PIs: A Feder, RH Pietrzak, SM Southwick

### Author Declarations

The study, conducted between April 2013 and September 2017, was approved by the Institutional Review Board of the Icahn School of Medicine at Mount Sinai, and all participants provided written informed consent.

### Summary of Updates

Updated text and added references.

